# Comparing Methods to Identify which Adult Emergency Department Visits Might be Avoided: A Retrospective Analysis of the MIMIC-IV-ED Database

**DOI:** 10.1101/2025.11.12.25340098

**Authors:** J.G. Wrightson, L.K. Truong, C. Starcevich, P. Truter, K.M. Khan, J. Morgan, K. McGrail, C.L. Ardern

## Abstract

**Background:** There are multiple ways to define and identify potentially avoidable emergency department visits, making it difficult to estimate their frequency accurately. In this study, we compared the proportions and characteristics of potentially avoidable ED visits identified using three commonly used algorithms.

**Methods:** We analyzed the publicly available Medical Information Mart for Intensive Care IV Emergency Department dataset (MIMIC-ED) to estimate the proportions and characteristics of potentially avoidable ED visits identified using i) the eventual discharge diagnosis (*Diagnosis*), ii) the use of hospital resources (*Resources*), and iii) the triage acuity score assigned to the patient during emergency department triage (*Triage-Acuity*). We found that the proportions and characteristics of potentially avoidable ED visits were affected by the algorithm used to identify them.

**Results:** The proportion of visits identified as potentially avoidable differed significantly between algorithms (2 - 21%), and few visits (<2%) were identified as potentially avoidable by all three algorithms. Presenting complaints, discharge diagnoses, and patient demographics were all similarly affected.

**Conclusion:** Methods for identifying potentially avoidable ED visits are not interchangeable. Choosing the appropriate definition and classification method will require researchers to carefully consider the types of visits and the characteristics of patients they wish to identify.

## Background

Emergency Departments (EDs) are in crisis. Overcrowding challenges emergency staff who are trying to deliver effective and timely care in the ED [1, 2] and detrimentally impacts patient care [3]. It has been widely reported that many ED visits are by patients who do not require treatment in the ED [4–6]. Methods to redirect these patients to appropriate care pathways have been developed, including from the ED to primary care centers [7] or specialists embedded within the ED [8]. Designing and successfully implementing strategies to redirect patients requires accurate population-level estimates of how many, and which, patients can be redirected. Because there are multiple ways to identify ED visits that might be appropriate for redirection, it is difficult to accurately estimate their frequency [6, 9, 10].

ED visits by patients who may not require ED care have been described as “non-urgent” [11], “preventable” [12], “non-emergent” [13], “avoidable” [14], “inappropriate” [15], and “low-acuity” [9]. These terms are often used interchangeably, yet they reflect overlapping but distinct paradigms for classifying ED use, and may capture different types of ED visits and patients [14]. Importantly, they do not necessarily reflect the reasons why patients are motivated or required to attend the ED [16–18]. In this paper, we use the term “*potentially avoidable ED visits*” to emphasize the goal of reducing such presentations while ensuring that patients continue to receive timely, high-quality care.

There are multiple methods to classify potentially avoidable ED visits, predominantly using three different types of algorithm: i) diagnosis-based algorithms, which classify visits/patients according to the patient’s final diagnosis [19–21], ii) resource-use algorithms, which classify visits based on the hospital resources used, including patient admission status [12, 22, 23], and iii) triage-based algorithms, which use the urgency or severity assigned at the initial presentation, often quantified by a triage-acuity score [24–26]. The language used in the classification methods does not always align intuitively with the operational definition used for potentially avoidable ED visits. For example, researchers have identified “low-acuity” visits using a diagnosis-based algorithm [27] and “non-urgent visits” using an acuity-score-based algorithm [28].

Estimates for the number of potentially avoidable ED visits by adults have been made using the three different classification algorithms; however, these estimates have not been directly compared against each other [29–31]. We do not know whether the different definitions and classification methods used to identify ED visits affect the frequency, characteristics of visits, and patients that are identified as potentially avoidable. The aim of this study was to examine the effect of different classification algorithms on characterizing potentially avoidable ED visits. We examined how different methods to identify ED visits could affect estimates of how many and which patients and visits are appropriate to redirect from the ED.

## Method

This study was a cross-sectional retrospective analysis of a publicly available administrative electronic health record dataset. The study received institutional ethics approval from The University of British Columbia (H25-01308). Reporting followed the REporting of studies Conducted using Observational Routinely-collected health Data (RECORD) guidelines [32].

### Data Sources and Participants

The analysis was performed using version 2.2 of the Medical Information Mart for Intensive Care IV Emergency Department (MIMIC-ED) dataset [33, 34], a dataset of ∼425,000 de-identified admissions to the Beth Israel Deaconess Medical Center ED (Boston, USA) collected between 2011 and 2019. MIMIC-ED contains data from patients admitted to the hospital from the ED, patients discharged home after treatment in the ED, and patients who left the hospital voluntarily before receiving care in the ED. The *edstays, diagnosis*, and *triage* tables from the MIMIC-ED *ed* module were joined using the *subject_id* and *stay_id* variables. We linked data from MIMIC-ED to the *patients* table from the MIMIC-IV *hosp* module using the *subject_id* variable.

We included adults (age at least 18 years) in this study: visits where participants’ age was <18 years (n = 4372) or not recorded (n = 76) were removed. Visits without an assigned triage acuity score (n = 6934) were removed. Primary diagnoses were taken from the ICD-9 or ICD-10 diagnosis assigned in the ED, or if the patient was admitted, in the hospital. We performed analyses with a single visit per patient. Where patients had multiple visits, we included the first visit in our analysis. Data processing steps can be found in the online materials.

### Algorithms to Identify Potentially Avoidable ED Visits

We compared three common algorithms for identifying potentially avoidable ED visits:

1. A diagnosis-based algorithm (“*Diagnosis*”) commonly referred to as the New York University Emergency Department algorithm [35, 36]. Full details of the algorithm can be found elsewhere [37]. Briefly, the algorithm assigns probabilities to the ICD (9 and 10) primary diagnosis codes across four categories: Non-emergent, Emergent-Primary Care, and two Emergent-ED categories. We labelled a visit as potentially avoidable if the summed probability for the Non-emergent and Emergent-Primary Care categories was over 0.5 [37].
2. A resource-based algorithm (“*Resources*”), which uses events that occurred during the ED visit and the disposition (discharge location) [23, 38]. Visits where patients were not admitted to the hospital, were not kept in the ED under observation, were discharged home, and did not receive medications while in the ED, were labelled as potentially avoidable.
3. A triage-acuity algorithm (“*Triage-Acuity*”), which uses the triage-acuity score assigned to the patient on arrival to the ED. Triage acuity is recorded in MIMIC-ED using the Emergency Severity Index (ESI) acuity scale, where a higher score represents less acute patients/visits. Visits where patients were assigned either a 4 or 5 on the ESI at triage were labelled as potentially avoidable [39].

### Outcomes

We examined the effect of each classification algorithm on the following outcomes: proportion of ED visits (%) identified as potentially avoidable, most common presenting complaint, three most common discharge diagnoses (rank), patient age (years), sex (female/male, labeled as “gender” in MIMIC-ED), self-reported race, self-reported pain score at triage (/10), acuity assigned at triage (ESI, /5), discharge disposition (Home/Admitted/Other), and length of stay in the ED (minutes) of potentially avoidable ED visits.

### Analysis

Analyses were performed using R (version 4.5.0) [40] and Python (version 3.12.2) [41]. Full analysis details, including a list of the packages and libraries used for analysis, can be found in the online materials. Summary statistics are reported as either mean [95%CI], median [IQR], or proportion [95%CI]. Confidence intervals for proportions were calculated using the Agresti-Coull interval. We used an UpSet plot [42] and Cohen’s Kappa to identify the intersections and agreements between the potentially avoidable visits identified using each algorithm. For the free-text within ‘presenting complaint’, we generated sentence embeddings using a BioBERT sentence-transformer [43], which were then reduced with principal component analysis (80 components) and clustered using k-means clustering (k = 12). Representative n-grams were extracted from each cluster and summarized for interpretability. Conditions were extracted from the ICD-10-CM and ICD-9-CM chapters for the common primary diagnoses identified by each algorithm.

## Results

We analyzed data from 201,106 visits to the ED that occurred between 2011 and 2019. Patient demographics for all ED visits and the potentially avoidable visits identified with the three algorithms are shown in Table 1.

**Table 1.**
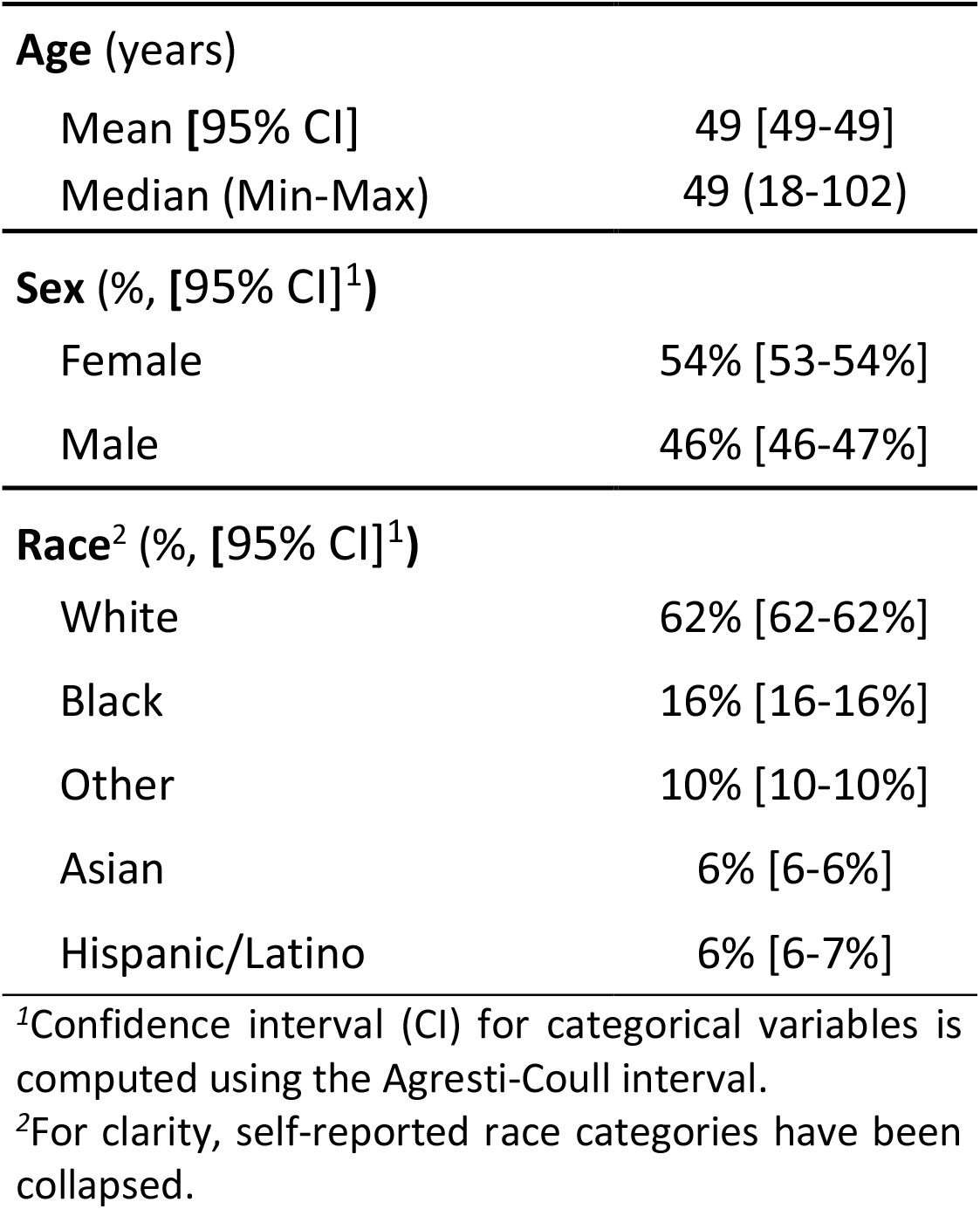
Patient demographics.

The numbers of potentially avoidable ED visits that were identified by each algorithm are shown in Figure 1. There were 100,568 visits (≈50% of all ED visits) identified as potentially avoidable by at least one classification algorithm. The proportions and patient and visit characteristics for the potentially avoidable ED visits are shown in Table 2. Few visits (≈2% of all ED visits) were identified as potentially avoidable by all three algorithms. The *Diagnosis* algorithm identified the largest number of unique visits and had the largest proportion of visits that resulted in admission to the hospital. The Cohen’s Kappa values for agreement were small, suggesting that agreement was poor between the algorithms (*Diagnosis* x *Resources* = 0.1 [0.1-0.1], *Diagnosis* x *Triage-Acuity* < 0.1 [0.0-0.0], *Resources* x *Triage-Acuity* = 0.1 [0.1-0.1]).

**Table 2.** Proportions and characteristics of visits and patients, for all ED visits and visits uniquely identified by each algorithm.

|  | All ED visits | Potentially Avoidable ED Visits (by Algorithm) |  |  |  |  |  |  |
| --- | --- | --- | --- | --- | --- | --- | --- | --- |
|  |  | Diagnosis | Resource | Triage Acuity | Diagnosis+<br>Resources | Resources+<br>Triage Acuity | Diagnosis+<br>Triage Acuity | Diagnosis+<br>Resources+<br>Triage Acuity |
| **Number of visits**<br>(N unique visits) | 201106 | 44119 | 23427 | 4690 | 17306 | 4278 | 3578 | 3170 |
| **Proportion**<br>(% [95%CI]<sup>1</sup>) | 100<br>[100-100] | 21.9<br>[21.8-22.1] | 11.6<br>[11.5-11.8] | 2.3<br>[2.3-2.4] | 8.6<br>[8.5-8.7] | 2.1<br>[2.1-2.2] | 1.8<br>[1.7-1.8] | 1.6<br>[1.5-1.6] |
| **Age**<br>(Years, mean [95%CI]) | 49.2<br>[49.1-49.3] | 47.0<br>[46.9-47.2] | 42.2<br>[42.0-42.5] | 38.1<br>[37.6-38.5] | 44.5<br>[44.2-44.8] | 35.3<br>[34.9-35.8] | 37.8<br>[37.3-38.4] | 37.3<br>[36.7-37.9] |
| **Female**<sup>2</sup><br>(%, [95%CI]<sup>1</sup>) | 53.6<br>[53.4-53.8] | 60.9<br>[60.4-61.4] | 50.1<br>[49.4-50.7] | 50.3<br>[48.9-51.8] | 59.5<br>[58.7-60.2] | 49.5<br>[48.0-51.0] | 57.4<br>[55.7-59.0] | 52.0<br>[50.2-53.7] |
| **Pain**<br>(/10, Median, [IQR]) | 4<br>[0-7] | 6<br>[2-8] | 0<br>[0-5] | 5<br>[2-8] | 4<br>[0-7] | 4<br>[0-6] | 7<br>[4-8] | 4<br>[0-7] |
| **Acuity**<sup>3</sup><br>(/5, Median, [IQR]) | 3<br>[2-3] | 3<br>[2-3] | 3<br>[2-3] | 4<br>[4-4] | 3<br>[3-3] | 4<br>[4-4] | 4<br>[4-4] | 4<br>[4-4] |
| **Length of stay**<br>(Mins, mean [95%CI]) | 392<br>[390-393] | 414<br>[411-417] | 297<br>[294-300] | 214<br>[209-219] | 261<br>[259-264] | 157<br>[154-160] | 213<br>[207-219] | 151<br>[148-155] |
| **Discharged Home**<br>(%, [95%CI]<sup>1</sup>) | 60.4<br>[60.2-61.0] | 74.9<br>[74.5-75.3] | 91.4<br>[91.0-91.8] | 95.3<br>[94.6-95.8] | 86.6<br>[86.1-87.1] | 95.4<br>[94.8-96.0] | 96.7<br>[96.1-97.2] | 87.0<br>[85.8-88.1] |
| **Admitted to Hospital**<br>(%, [95%CI]<sup>1</sup>) | 34.2<br>[34.0-34.4] | 22.6<br>[22.2-23.0] | 0.0<br>[0.0-0.0] | 3.6<br>[3.1-4.2] | 0.0<br>[0.0-0.0] | 0.0<br>[0.0-0.0] | 1.5<br>[1.1-1.9] | 0.0<br>[0.0-0.0] |

<sup>1</sup> Confidence interval (CI) computed using Agresti-Coull interval<sup>2</sup> Only female/male are reported in the data<sup>3</sup> Emergency Severity Index, 1 = most severe

**Figure 1.**
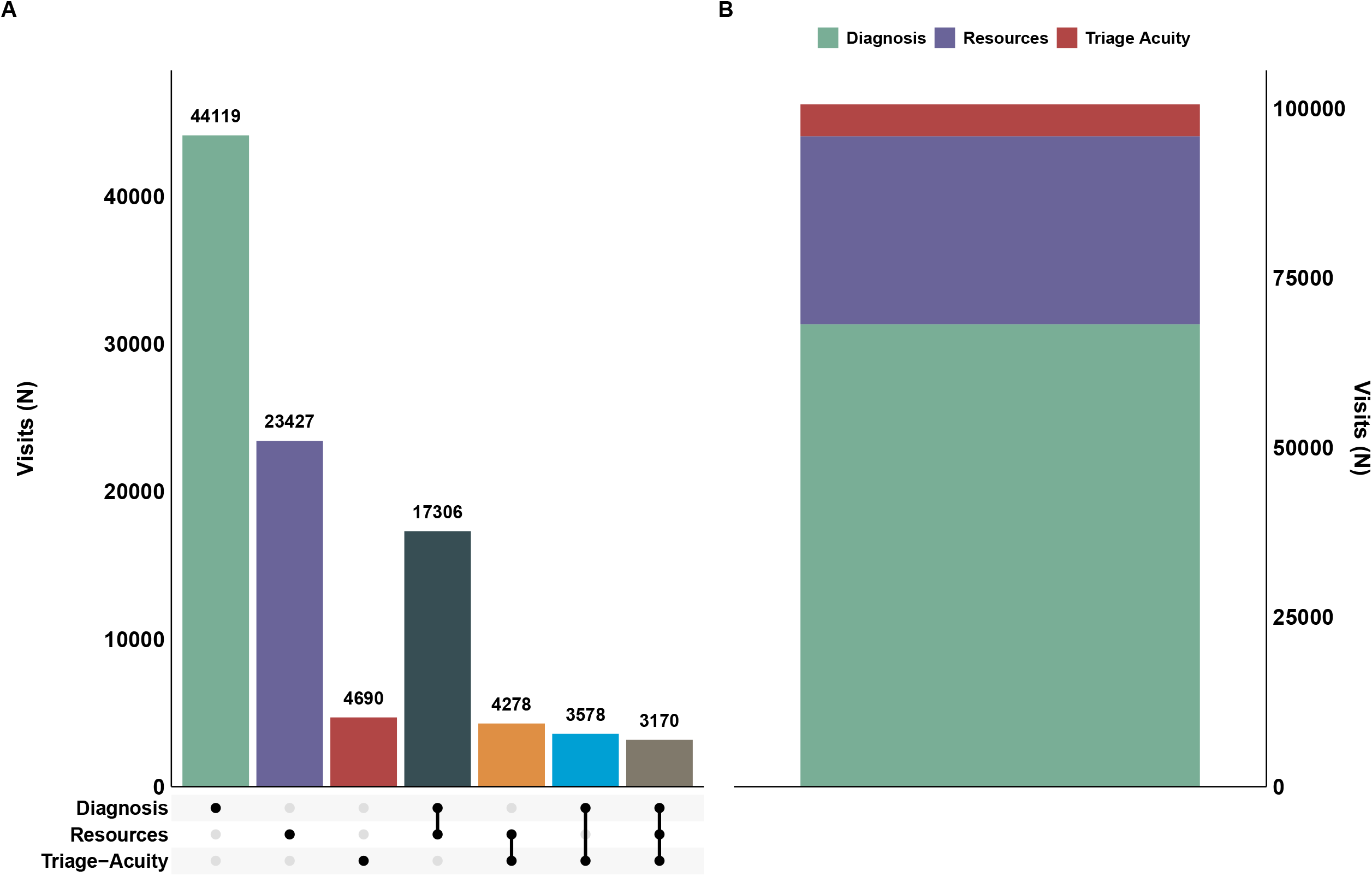
The counts and intersections between the potentially avoidable ED visits identified by each classification algorithm. Panel A (left) shows the intersection of the visits identified with individual and combinations of algorithms. The individual and combinations of algorithms are indicated by single and joined closed circles in the matrix, respectively. The bar displays the number of visits uniquely identified by individual and combinations of algorithms identified in the matrix. Data are only for visits that were identified by at least one algorithm. Panel B (right) shows the cumulative counts of potentially avoidable visits identified by each algorithm.

Common presenting complaints and conditions represented in the potentially avoidable ED visits are shown in Table 3 and Table 4. There were no chief complaints or diagnoses that were among the most commonly identified by all three algorithms.

**Table 3.** Common presenting complaints for visits identified by each algorithm^1^.

| Rank | All ED visits | Potentially Avoidable ED Visits (by algorithm, % of visits) |  |  |  |  |  |  |
| --- | --- | --- | --- | --- | --- | --- | --- | --- |
|  |  | Diagnosis | Resources | Triage Acuity | Diagnosis+<br>Resources | Resources+<br>Triage Acuity | Diagnosis+<br>Triage Acuity | Diagnosis+<br>Resources+<br>Triage Acuity |
| 1st | Wound<br>evaluation<br>(21.0%) | Abdomen pain<br>(19.1%) | Wound<br>evaluation<br>(22.8%) | Wound<br>evaluation<br>(36.1%) | Abdomen pain<br>(18.4%) | Wound<br>evaluation<br>(41.3%) | Lower back<br>pain (31.5%) | Vehicle<br>collision +<br>Altered mental<br>status (21%) |
| 2nd | Vehicle<br>collision +<br>altered mental<br>status (12.1%) | Lower back<br>pain (12.4%) | Chest pain +<br>dyspnea<br>(14.8%) | Leg + knee pain<br>(26.2%) | Wound<br>evaluation<br>(14.8%) | Leg + knee pain<br>(26.1%) | Leg + knee pain<br>(16.2%) | Wound<br>evaluation<br>(20.1%) |
| 3rd | Abdomen pain<br>(12.1%) | Vehicle<br>collision +<br>Altered mental<br>status (12.3%) | Vehicle<br>collision +<br>Altered mental<br>status (14.2%) | Lower back<br>pain (16.6%) | Vehicle<br>collision +<br>Altered mental<br>status (13.7%) | Vehicle<br>collision +<br>Altered mental<br>status (12.8%) | Influenza like<br>illness (14.9%) | Influenza like<br>illness (18.2%) |

<sup>1</sup> From cluster analysis of free-text presenting complaints (see method)

**Table 4.** Common diagnosed conditions for visits identified by each algorithm^1^.

| Rank | All ED visits | Potentially Avoidable ED Visits (by algorithm, % of ED visits) |  |  |  |  |  |  |
| --- | --- | --- | --- | --- | --- | --- | --- | --- |
|  |  | Diagnosis | Resources | Triage Acuity | Diagnosis+<br>Resources | Resources+<br>Triage Acuity | Diagnosis+<br>Triage Acuity | Diagnosis+<br>Resources+<br>Triage Acuity |
| 1st | Non-specific<br>diagnoses<br>(23.1%) | Ill-defined<br>conditions<br>(44.6%) | Injury and<br>poisoning<br>(31.5%) | Injury and<br>poisoning<br>(82.8%) | Ill-defined<br>conditions<br>(51.6%) | Injury and<br>poisoning<br>(80.3%) | Diseases of the<br>MSK system<br>(43%) | Diseases of the<br>MSK system<br>(29.6%) |
| 2nd | Injury and<br>poisoning<br>(21.2%) | Diseases of the<br>MSK system<br>(15.3%) | Ill-defined<br>conditions<br>(27.5%) | Diseases of the<br>MSK system<br>(4%) | Diseases of the<br>MSK system<br>(16.8%) | Mental<br>disorders<br>(3.8%) | Diseases of the<br>skin and<br>subcutaneous<br>tissue (10.5%) | Ill-defined<br>conditions<br>(16.8%) |
| 3rd | Diseases of the<br>circulatory<br>system (7.9%) | Diseases of the<br>skin and<br>subcutaneous<br>tissue (7.7%) | Mental<br>disorders<br>(17.6%) | Factors<br>influencing<br>health<br>status/contact<br>with health<br>services (2.3%) | Diseases of the<br>genitourinary<br>system (5.3%) | Factors<br>influencing<br>health<br>status/contact<br>with health<br>services (3.2%) | Ill-defined<br>conditions<br>(10.3%) | Diseases of the<br>skin and<br>subcutaneous<br>tissue (10.4%) |

MSK; Musculoskeletal
<sup>1</sup> From the ICD-9-CM and ICD-10-CM chapters for the primary diagnosis ICD code
Non-specific diagnoses = ICD-9: 780-789, ICD-10: R00-R99
Injury and poisoning = ICD-9: 800-999, ICD-10: S00-T88
Mental, behavioral & neurodev. disorders = ICD-9: 290-319, ICD-10: F00-F99
Diseases of the circulatory system = ICD-9: 390-459, ICD-10: I00-I99
Diseases of the MSK system = ICD-9: 710-739, ICD-10: M00-M99
Diseases of the skin and subcutaneous tissue = ICD-9: 680-709, ICD-10: L00-L99
Factors influencing health status/contact with health services = ICD-9: V01-V91, ICD-10: Z00-Z99

## Discussion

Our results suggest that methods for identifying potentially avoidable ED visits are not interchangeable: the choice of the algorithm determines how many and which patients and conditions are identified as potentially avoidable. There was little agreement between the different methods used to identify potentially avoidable ED visits, and few visits were identified as potentially avoidable by all three classification algorithms. There were differences between the algorithms for the numbers and proportions of visits, the characteristics of the visits, and patients. Choosing the appropriate definition and classification method will require researchers to carefully consider the types of visits and the characteristics of patients they wish to identify.

Our results extend previous findings [29–31] by showing that classification methods not only influence estimates of the frequency and proportion of potentially avoidable ED visits, but also affect the types of patients, conditions, and visits identified as potentially avoidable. The *Diagnosis* algorithm identified the greatest number of potentially avoidable ED visits. The greatest overlap in identified visits occurred between the *Diagnosis* and *Resources* algorithms, but even this intersection was smaller than the number of visits identified by either algorithm individually. In comparison, few visits were identified solely by the *Triage-Acuity* algorithm.. Accurate estimates of the number of patients or visits, and the type of care are essential for effective health care planning and health system needs assessments [44]. Although there is increasing interest in developing methods to reduce potentially avoidable ED visits, the strength of the evidence supporting these approaches is unclear, in part because of the heterogeneity in how the visits are defined and identified [9, 10].

In our analysis, the median triage acuity score for potentially avoidable visits identified with either the *Diagnosis* or *Resources* algorithms (3/5) was higher than the score (4/5) typically used as a cut-off for the *Triage-Acuity* algorithm. Compared to using *Diagnosis* or *Resource-based* methods, there is little benefit in using *Triage-Acuity* to retrospectively identify potentially avoidable ED visits. The relatively large disparities between the numbers of unique and shared visits identified by the *Diagnosis* and *Resource-*based algorithms indicate that one cannot assume the estimates for the numbers of potentially avoidable ED visits identified with one algorithm represent the numbers of visits identified by the other.

The choice of algorithm, in effect, determines the frequency and types of patients, conditions, and visits that could be considered potentially avoidable. The definitions are not interchangeable, and we suggest that the validity of the definitions is uncertain. This is important because the choice of algorithm has direct implications for healthcare planning and resource use. For instance, patients identified by the three algorithms presented with different complaints, reported varying pain levels at triage, received distinct diagnoses, and were hospitalized in differing numbers. In addition, although the *Diagnosis* algorithm identified more visits than the other algorithms, the *Triage-Acuity* does not identify some visits for injuries and poisoning that could be redirected from the ED, and may underestimate the number of potentially avoidable visits. Researchers, health care administrators, and policy makers should carefully consider the definitions and assumptions underlying each algorithm when selecting one to identify potentially avoidable ED visits. When attempting to identify the types of services that can be implemented and the resources needed for effective delivery, researchers must carefully consider the implications of the algorithm used to identify potentially avoidable ED visits. We encourage researchers to consider which patients, conditions, and visits they consider “less urgent” than others, and select an algorithm that best identifies those visits.

It has long been suspected that the criteria used to define and identify potentially avoidable ED visits may not be interchangeable [29–31]. One solution to the disparity in definitions and methods used to identify potentially avoidable ED visits is to develop a gold standard case definition [14, 38]. This approach assumes there is a true class of ED visits that are potentially avoidable/non-urgent/non-emergent. An alternative, but less widely adopted, approach is to adopt a utilitarian perspective and assume that the disparity between algorithms has utility. With this approach, researchers and health administrators would treat each algorithm as a means of classifying different types of potentially avoidable ED visits and patients. For example, if the primary purpose is to measure the frequency of patients with certain conditions who could be redirected from the ED, a diagnosis-based algorithm might be more suitable than if the purpose were to identify patients who would not be admitted. If both are important, then combining algorithms may be preferable to, for example, identifying patients with pre-defined conditions who are less likely to use significant hospital resources. One limitation of both the *Diagnosis-* and *Resource-based* algorithms is that they can only be used retrospectively. If the aim is to identify visits that can be redirected before attending the ED or from within the ED, then more advanced algorithms must be used [45, 46]

A limitation of our study, and more generally of the oft-used definitions for potentially avoidable ED visits, is that they do not reflect the complexity of ED visits and clinical presentations [47]. For example, a 90-year-old patient with a musculoskeletal condition classified as potentially avoidable by the *Diagnosis* algorithm may still require ED care, whereas a 20-year-old with the same condition may not. Similarly, visits by patients diagnosed with mental, behavioural, or neurodevelopmental disorders (one of the most common diagnoses identified by the *Resource* algorithm) may not use many ED resources, but attending the ED may be a necessary first step to accessing care. We encourage researchers and administrators to consider a priori which conditions and patients should *not* be classified as potentially avoidable, and refine their choice of algorithm accordingly. Another limitation is that the data are from a single center. It is not clear whether the characteristics of patients and visits reported here would generalize to other health systems (especially those that use other triage acuity scales) and to pediatric patients. However, we expect the disparity between algorithms to be evident in similar healthcare systems (e.g., [29–31]).

## Conclusion

The choice of algorithm influenced the number and types of visits and patients that were identified as potentially avoidable ED visits. Rather than using a single algorithm to identify potentially avoidable ED visits, we encourage researchers and health care systems to develop or refine algorithms that reflect the purpose and priorities of their own analysis. We encourage analysts to adopt a utilitarian view of the disparities between different algorithms by contrasting results derived from different algorithms.

## Data Availability

The MIMIC-ED data are publicly available from Physionet [48]. Code is available online: https://github.com/James-G-Wrightson/mimiced_compare.git

## Notes

### Competing Interest Statement

The authors have declared no competing interest.

### Funding Statement

This study did not receive any funding

### Author Declarations

The MIMIC-ED data are publicly available from Physionet (https://physionet.org/content/mimic-iv-ed/2.2/)

